# Pilot Randomized Trial of Two Food Purchasing Interventions in Hypertensive Individuals in New Orleans: An American Heart Association Food is Medicine Initiative

**DOI:** 10.1101/2025.08.01.25332844

**Authors:** Manasi Tannu, Rebecca Young, Amanda Brucker, Bridget Simon-Friedt, Yuriy Bisyuk, Sylvester Tumusiime, Lindsey Rudov, Jessica Stein, Meagan Alley, Darcy Hannagan, Madeline Young, Jasmine McGary, Sarah Palmer, Lauren Cohen, Neha Pagidipati, Adrian Hernandez, Thomas W. Carton

## Abstract

**Background:** Consuming fruits and vegetables daily is linked to improved health in individuals with hypertension, yet access to nutritious foods remains limited. Voucher incentives have been shown to improve healthy food purchasing.

**Objective:** Compare the effects of an online versus in-store voucher intervention on food purchasing and health outcomes in hypertensive individuals.

**Methods:** In this randomized, pilot trial, 98 participants with uncontrolled hypertension received a $100 monthly food voucher to either an online grocery platform or an in-store local grocery. Both groups received educational materials. The primary outcome was change in self-reported fruit and vegetable consumption. Secondary outcomes were voucher utilization, engagement with educational materials, and changes in BP, weight, and BMI. Generalized estimating equations for repeated measures were used to assess changes in food-group consumption over the five study months.

**Results:** There was no statistically significant difference between arms for self-reported fruit and vegetable consumption. There was an overall 20% (IQR -17%, 93%) increase in fruit and vegetable consumption within both arms. Voucher utilization was significantly higher in the in-store arm (>90%) than the online arm (<64%). Engagement with educational materials was greater in the in-store arm. No significant changes in weight, BMI or BP were observed.

**Conclusion:** There was no significant difference between in-store and online vouchers for fruit and vegetable consumption. Higher engagement and voucher utilization were observed in the in-store arm than the online arm. Future interventions should address technological barriers and be conducted in larger populations to enhance program effectiveness and assess long-term health impacts.

## INTRODUCTION

Poor dietary habits are strongly associated with adverse health outcomes, including cardiovascular disease, diabetes, and colorectal cancer.^1^ Evidence increasingly supports the role of healthy food programs in improving clinical markers such as blood pressure (BP), glycemic control, and body mass index (BMI).^2–4^ In recognition of the potential for nutrition-based interventions to enhance health outcomes, the American Heart Association (AHA) launched the *Health Care by Food* initiative in 2024. This initiative has funded 20 Food is Medicine (FIM) pilot studies aimed at evaluating the scalability, cost-effectiveness, and implementation challenges of nutrition-focused healthcare strategies.^5^

Current dietary guidelines from the U.S. Departments of Health and Human Services (DHHS) and Agriculture (USDA) recommend consuming 4–5 servings of fruits and vegetables daily to lower the risk of mortality.^3,4^ Prospective meta-analyses have further demonstrated that higher fruit and vegetable consumption is associated with reduced risk of hypertension.^6^ However, many individuals face structural and financial barriers to purchasing fresh produce, including food insecurity, limited grocery access, and economic instability.

Voucher-based financial incentives—coupons redeemable for nutritious foods—have been shown to improve healthy food purchasing and clinical outcomes such as diabetes and hypertension, making them a promising strategy for food-insecure populations with hypertension.^2,7,8^ Additionally, the expansion of online grocery stores and rapid delivery platforms presents new opportunities to improve food access by reducing geographical barriers to healthy food options. However, the effectiveness of these digital solutions remains understudied, requiring targeted interventional research.

This pilot, randomized study sought to compare the feasibility and effectiveness of two voucher-based interventions —online versus in-store—on food purchasing and consumption behaviors among individuals with hypertension. The study evaluated (1) changes in food purchasing and consumption patterns in the overall cohort, (2) the impact of in-store versus online voucher use on healthy food purchasing and consumption, (3) changes in weight, BMI, and BP over five months in each group and overall, and (4) barriers to implementing voucher-based programs.

## METHODS

### Study Population

Patients were recruited from the University Medical Center (UMCNO), New Orleans, LA, a partner health system within the Research Action for Health Network (REACHnet), which is a Clinical Research Network (CRN) within PCORnet^®9^. Electronic health records (EHR) were queried via the PCORnet^®^ Common Data Model to identify eligible participants. Participants who met inclusion criteria were ≥50 years of age, with uncontrolled hypertension (defined as at least two documented blood pressure [BP] readings of systolic >140 mmHg or diastolic >90 mmHg within the prior 6 months), had a working phone number or valid email address, and were English-speaking. Exclusion criteria included an absence of internet access (either at home, library, relative or through phone plan), lack of a secondary form of electronic payment (credit card, debit card, SNAP, EBT) to establish an online grocery account, and/or no adequate transportation to access a grocery store (ie. no personal car, reliable ride, access to public transportation or ride-hailing service) in the New Orleans area.

Study coordinators first contacted eligible participants through MyChart or REDCap via an electronic message that included a link to the HUGO platform, an online platform that facilitates data collection and engagement with study participants.

Baseline demographic and clinical variables such as age, blood pressure (BP), and body mass index (BMI) were derived from the UMCNO electronic health records. Comorbidities were derived using diagnosis ICD-10 codes from participants’ EHR from the year prior to their baseline and publicly available Elixhauser comorbidity software.^10^ Participants self-reported demographic characteristics such as household income, highest level of education achieved, receipt of SNAP benefits, employment status, health insurance type, and number of people in household at baseline via an enrollment survey.

### Study design and interventions

This study was a randomized, multi-arm parallel group design, with an allocation ratio of 1:1 in two intervention arms: in-store versus online grocery. Participants were assigned to either arm following a block randomization scheme with a block size of four. Randomization was performed within a REDCap randomization module. Because the study was designed as a pilot, it was not powered to detect statistical differences. The enrollment target was N=100 participants, with n=50 in each arm.

Participants were assigned to one of two interventions: (1) a voucher delivered through an online grocery delivery platform or (2) a voucher of equal value usable at a large, local supermarket chain in New Orleans. Vouchers were $100 per month for 5 months, regardless of intervention arm. For the online arm, delivery fees were waived but local service fees were not. While online vouchers could be restricted to exclude purchases of alcohol and tobacco, such restrictions were not feasible for in-store vouchers. No additional restrictions could be applied to either voucher type, allowing participants to use them for non-food items. Step-by-step guides were provided for both online and in-store voucher usage and participants were educated and encouraged to use the vouchers only for food items, specifically for fruits and vegetables. Technical assistance was provided by the study team.

Study procedures were reviewed and approved by the Duke University School of Medicine Institutional Review Board (Pro00114628). This work was supported by AHA Health Care x Food initiative, grant 24RPGFIM1198185 / Duke University / 2023.

### Educational Materials

The educational intervention was designed as a self-directed intervention. Educational materials were developed by the research team in collaboration with Health in Our Hands (HiOH), a PCORnet^®^ affiliated patient advisory group. Topics suggested by HiOH patients were then synthesized and curated into a set of educational fact sheets covering topics such as shopping for healthy food on a budget, interpreting nutritional labels, preparing quick and healthy meals, and understanding the nutritional value of frozen/canned foods. These resources were stored on a study web page which was sent to participants each month via email. Participant utilization of educational materials was assessed by self-reported survey.

### Food Consumption Data

At the end of each month, participants were asked to complete an online Food Frequency Questionnaire (FFQ) in which they self-reported their average weekly consumption of fruits (excluding fruit juice), vegetables (salad, vegetables, and potatoes), dairy (milk, cheese, and yogurt), meat/seafood (poultry, red meat, and seafood), desserts/sweets (ice cream, dessert, sugary beverages, juice), and restaurant and fast food meals over the past 30 days as well as self-reported health status and physical activity. The FFQ was an abridged version of the National Health and Nutrition Examination Survey (NHANES) Dietary Screener Questionnaire.^11^ Questions were reviewed by a cardiologist and registered dietician. Self-reported consumption data were converted to number of servings per month for analysis. The surveys automatically expired after 7 days to prevent reporting overlap between months. FFQ surveys were administered at baseline and months 1-5 to capture longitudinal data on participant food consumption patterns. Participants received a financial incentive to complete surveys; $15 per completed survey and an additional $25 at study end if all surveys were completed.

### Purchase Data Categorization

Purchasing data were acquired from grocer partners and standardized to be consistent across platforms. Four research team members conducted an iterative qualitative assessment to map grocery data to a reduced set of FFQ categories (ie. fruits, vegetables, meat, dairy, sweets) for both grocery data platforms to achieve a consistent set of variables across arms.

### Statistical Analysis

The primary outcome was change in self-reported consumption of fruits and vegetables (combined) between baseline and 5 months. Secondary outcomes were: (1) changes in self-reported consumption in fruits, vegetables, sweets, meat, and dairy; (2) between-arm differences in voucher utilization at least once during each month; and (3) between-arm differences in the total dollar amount of voucher used. Additional exploratory outcomes were engagement with educational materials, and changes in BP, BMI, and weight between baseline and the final follow-up timepoint.

Baseline patient characteristics are presented for the overall study cohort and each study arm. Categorial variables are reported as counts (percentages) and continuous variables as medians (Q1, Q3). Between-arm standardized differences were calculated. For repeated measures analyses of self-reported consumption , last observation carried forward (LOCF) imputation method was employed to impute missing values; if a participant had a missing value at 5 months and a known value at 4 months, the 4-month value was used to impute the 5-month value. A generalized estimating equations (GEE) framework was used to estimate between-arm and within-arm changes in consumption between each month of data, utilizing an identity link and exchangeable working correlation structure. A weighted GEE framework was applied to estimate changes in BP, BMI, and weight while accounting for missingness in 5-month measurements. All GEE frameworks adjusted for a pre-specified set of covariates: age, household income category, SNAP benefits (yes/no), number of people in household (continuous), sex, and race. Model-estimated mean differences are reported, in addition to 95% confidence intervals derived from a robust standard error estimate and corresponding p-values.^12^

Voucher usage rates are reported by arm by month. A chi-squared test was implemented to test for differences in usage rates between the two arms. Median (IQR) amount of benefit spent is additionally reported by month and by arm, and a Kruskal Wallis test was utilized to test for differences. Rates of self-reported usage of educational materials were analyzed using a chi-squared test. The total amount of each voucher spent and the amount used to purchase fruits and vegetables are reported as median (IQR) for the online arm.

For all outcomes, if a participant dropped out of the study, only the months of study participation were analyzed. Statistical analyses were conducted in SAS 9.4 (SAS Institute Inc) and R version 4.3.2. Results were interpreted using a significance level of alpha = 0.05. Given the exploratory nature of the study, no multiple testing adjustments were made.

## RESULTS

### Baseline Characteristics

A total of 3,997 patients met inclusion criteria within the UMCNO EHR (**Figure 1 Consort diagram**) but 1,773 patients were excluded because they did not have a MyChart account. Of those, 98 were consented, enrolled, and completed baseline surveys. Participants were randomized to receive vouchers in either the in-store arm (n=49) or the online arm (n=49). One participant self-withdrew from the online arm before data was collected.

**Figure 1.**
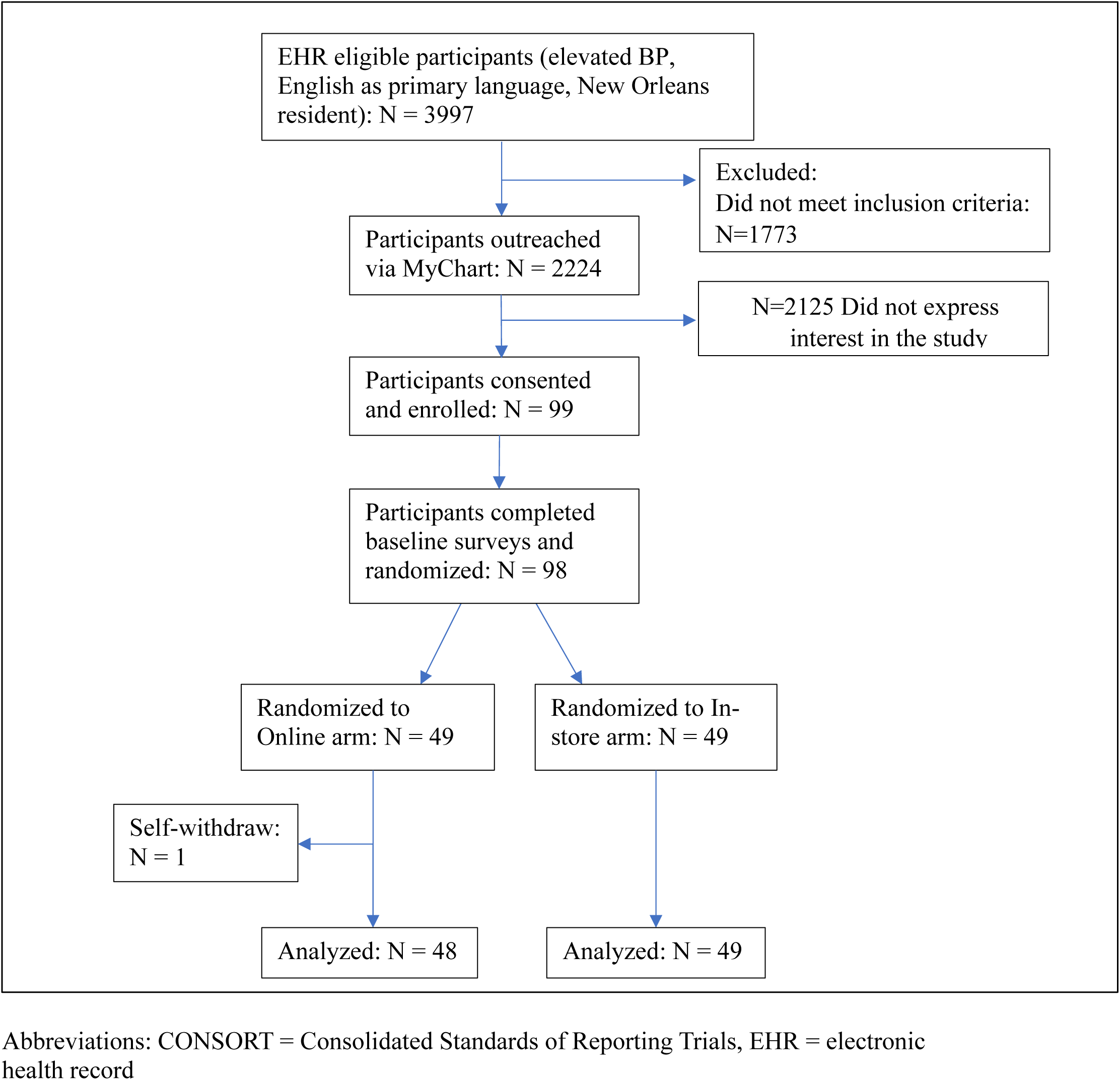
CONSORT diagram.

Baseline characteristics between the two arms are displayed in **Table 1**. Briefly, enrolled participants had a median age of 57 years (interquartile range [IQR] 53-63), 65% were female, 61% were Black, and 49% had an income less than $20,000. The baseline average systolic BP was 140.2 mmHg, diastolic BP was 81.5 mmHg, 23% had malignant hypertension, and 14% had peripheral vascular disease.

**Table 1.** Baseline Clinical and Demographic Characteristics.

|  | **Total**<br>**(N=97)** | **In-store**<br>**(N=49)** | **Online**<br>**(N=48)** | **Standardized**<br>**Difference\*** |
| --- | --- | --- | --- | --- |
| **Demographics** |  |  |  |  |
| Age (median, IQR) | 56.5<br>(53, 63) | 56.0<br>(53.0, 63.0) | 57.0<br>(53.0, 62.0) | 0.01 |
| Female (%) | 64.9 | 67.3 | 62.5 | 0.05 |
| Race (%) |  |  |  | 0.21 |
| Black | 60.8 | 57.1 | 64.6 |  |
| White | 28.9 | 34.7 | 22.9 |  |
| Multiple Races | 6.2 | 4.1 | 8.3 |  |
| Prefer not to Answer | 4.1 | 4.1 | 4.2 |  |
| Ethnicity (%) |  |  |  | 0.02 |
| Hispanic | 5.2 | 6.1 | 4.2 |  |
| Non-Hispanic | 86.6 | 85.7 | 87.5 |  |
| Prefer not to answer | 8.2 | 8.2 | 8.3 |  |
| Income Category (%) |  |  |  | 0.03 |
| High Income, >\$50,000 | 10.3 | 6.1 | 14.6 | |
| Middle Income, \$20,000-\$50,000 | 37.1 | 34.7 | 39.6 | |
| Low Income, <\$20,000 | 48.5 | 55.1 | 41.7 | |
| Education Level (%) |  |  |  | 0.02 |
| College Graduate | 26.8 | 26.5 | 27.1 |  |
| Some College | 48.5 | 49.0 | 47.9 |  |
| High School | 19.6 | 16.3 | 22.9 |  |
| No Diploma | 5.2 | 8.2 | 2.1 |  |
| SNAP Benefit Recipient (%) | 41.2 | 44.9 | 37.5 | 0.02 |
| Not working (%) | 63.9 | 59.2 | 68.8 | 0.28 |
| Insurance Type (%) |  |  |  | 0.18 |
| Private | 12.4 | 12.2 | 12.5 |  |
| Public (Medicare, Medicaid, VA) | 83.5 | 85.7 | 81.3 |  |
| Uninsured | 2.1 | 0 | 4.2 |  |
| Other | 2.1 | 2.0 | 2.1 |  |
| Number of people in household |  |  |  | 0.33 |
| 1 | 43.3 | 46.9 | 39.6 |  |
| 2 | 37.1 | 34.7 | 39.6 |  |
| 3 | 11.3 | 10.2 | 12.5 |  |
| ≥4 | 8.2 | 8.1 | 8.3 |  |
| BMI (kg/m<sup>2</sup>) (median, IQR) | 34.1<br>(28.0, 38.6) | 33.3<br>(27.0, 39.7) | 34.7<br>(30.6, 38.1) | 0.17 |
| Average SBP (mmHg) in prior year (median, IQR) | 140.2<br>(135.5, | 138.3<br>(134.7, | 140.9<br>(136.5, | 0.26 |
|  | 146.0) | 146.0) | 146.4) |  |
| Average DBP (mmHg) in prior year (median, IQR) | 81.5 (75.7, 87.3) | 81.2 (72.2, 86.8) | 82.8 (76.7, 88.3) | 0.16 |
| **Comorbidities (%)** |  |  |  |  |
| Malignant Hypertension | 22.7 | 14.3 | 31.3 | 0.42 |
| Non-malignant Hypertension | 66.0 | 53.1 | 79.2 | 0.62 |
| Cerebrovascular Disease | 3.1 | 0 | 6.3 | 0.38 |
| Peripheral vascular disease | 14.4 | 10.2 | 18.8 | 0.25 |
| Diabetes with end-organ damage | 23.7 | 18.4 | 29.2 | 0.26 |
| Diabetes without end-organ damage | 33.0 | 30.6 | 35.4 | 0.09 |
| Heart failure | 8.2 | 6.1 | 10.4 | 0.16 |
| Obesity | 37.1 | 30.6 | 43.8 | 0.27 |
| Chronic Lung Disease | 22.7 | 28.6 | 16.7 | 0.31 |

\*Standardized difference greater than 0.20 (corresponding to 20%) indicates a clinically meaningful difference between arms
Abbreviations: IQR = interquartile range, SBP = systolic blood pressure, DBP = diastolic blood pressure, SNAP = supplemental nutrition assistance program, HIV = human immunodeficiency virus, AIDS = acquired immunodeficiency syndrome, VA = veterans administration

### Change in Food Consumption Patterns between Randomization Arms

Self-reported food servings consumed per month in each arm are shown in **Table 2**. There was no significant difference between in-store or online vouchers for the primary outcome of combined fruit and vegetable consumption over 5 months of intervention (Adjusted mean difference in-store vs online: -1.13 servings per month, 95% CI [-12.56, 10.30]).

**Table 2.**
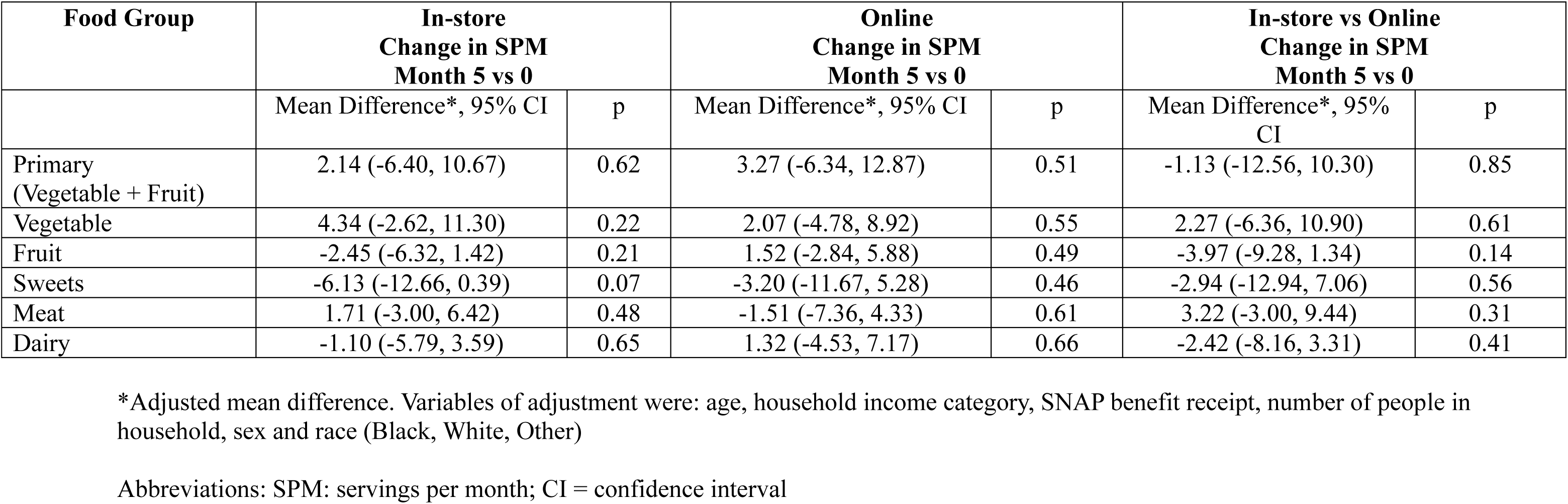
Adjusted Mean Differences in Servings per Month Stratified by Randomization Arm and Food Group.

| Food Group | In-store<br>Change in SPM<br>Month 5 vs 0 |  | Online<br>Change in SPM<br>Month 5 vs 0 |  | In-store vs Online<br>Change in SPM<br>Month 5 vs 0 |  |
| --- | --- | --- | --- | --- | --- | --- |
|  | Mean Difference*, 95% CI | p | Mean Difference*, 95% CI | p | Mean Difference*, 95% CI | p |
| Primary<br>(Vegetable + Fruit) | 2.14 (-6.40, 10.67) | 0.62 | 3.27 (-6.34, 12.87) | 0.51 | -1.13 (-12.56, 10.30) | 0.85 |
| Vegetable | 4.34 (-2.62, 11.30) | 0.22 | 2.07 (-4.78, 8.92) | 0.55 | 2.27 (-6.36, 10.90) | 0.61 |
| Fruit | -2.45 (-6.32, 1.42) | 0.21 | 1.52 (-2.84, 5.88) | 0.49 | -3.97 (-9.28, 1.34) | 0.14 |
| Sweets | -6.13 (-12.66, 0.39) | 0.07 | -3.20 (-11.67, 5.28) | 0.46 | -2.94 (-12.94, 7.06) | 0.56 |
| Meat | 1.71 (-3.00, 6.42) | 0.48 | -1.51 (-7.36, 4.33) | 0.61 | 3.22 (-3.00, 9.44) | 0.31 |
| Dairy | -1.10 (-5.79, 3.59) | 0.65 | 1.32 (-4.53, 7.17) | 0.66 | -2.42 (-8.16, 3.31) | 0.41 |

\*Adjusted mean difference. Variables of adjustment were: age, household income category, SNAP benefit receipt, number of people in household, sex and race (Black, White, Other)
Abbreviations: SPM: servings per month; CI = confidence interval

There was a trend towards an increase in vegetable consumption in both the in-store arm (month 5 vs month 0 mean difference: 4.34 servings/month, 95% CI [-2.62, 11.30]) and online arm (month 5 vs month 0 mean difference: 2.07 servings/month, 95% CI [-4.78, 8.92]) over 5 months of intervention, but these results did not reach statistical significance. Participants in the in-store arm self-reported a modestly greater increase in vegetable consumption than the online arm (adjusted mean difference in-store vs online: 2.27 servings per month, 95% CI [-6.36, 10.90]), but this did not meet statistical significance.

Similarly, there was an overall trend in both arms of decreasing sweets intake over 5 months of intervention with a trend towards a larger decrease in self-reported sweets consumption in the in-store arm (adjusted mean difference in-store vs online: -2.94 servings/month, 95% CI [-12.94, 7.06]), but these results were not statistically significant.

### Change in Clinical Outcomes after Intervention

There was no change in average weight, BMI, systolic BP, or diastolic BP after 5 months of intervention between the in-store or online arms in adjusted or unadjusted models (**Table 3**). After 5 months of intervention in both arms, there was a trend towards decreased weight (in-store month 5 vs month 0: -2.3 lbs [-8.7, 4.1] and online 5m vs baseline -4.8 lbs [-11.6, 2.1]) and systolic BP (in-store -4.1 mmHg [-10.5, 2.2] and online -3.5 mmHg [-10.5, 3.6]), but this did not reach statistical significance.

**Table 3.** Adjusted Mean Differences in Clinical Measures: within Arm and between Arms.

| Outcome | In-store<br>Month 5 vs 0 |  | Online<br>Month 5 vs 0 |  | In-store vs Online<br>Month 5 vs 0 |  |
| --- | --- | --- | --- | --- | --- | --- |
|  | Mean Difference*,<br>95% CI | p | Mean Difference*,<br>95% CI | p | Mean<br>Difference*,<br>95% CI | p |
| Weight (lbs) | -2.3 (-8.7, 4.1) | 0.49 | -4.8 (-11.6, 2.1) | 0.17 | 2.5 (-6.5, 11.5) | 0.58 |
| BMI (kg/m2) | -0.3 (-1.3, 0.8) | 0.64 | -0.5 (-1.5, 0.5) | 0.34 | 0.3 (-1.1, 1.6) | 0.72 |
| Systolic BP<br>(mmHg) | -4.1 (-10.5, 2.2) | 0.20 | -3.5 (-10.5, 3.6) | 0.33 | -0.7 (-10.1, 8.7) | 0.89 |
| Diastolic BP<br>(mmHg) | 0 (-4.5, 4.5) | 0.99 | 0.8 (-3.7, 5.3) | 0.73 | -0.8 (-7.0, 5.5) | 0.81 |

\*Adjusted mean difference. Variables of adjustment were: age, household income category, SNAP benefit receipt, number of people in household, sex and race (Black, White, Other)
Abbreviations: BP = blood pressure; lbs = pounds; BMI = body mass index; kg=kilograms; mmHg = millimeters of mercury; CI = confidence interval

### Voucher Utilization

There was significantly higher voucher utilization in the in-store arm than the online arm (p=0.0001) (**Table 4**) across all 5 months of intervention. Monthly voucher utilization in the in-store arm was 90-94% while utilization in the online arm was consistently less than 64%. There was no significant change in voucher utilization over time in either arm. Regarding voucher spending, participants in the in-store arm had consistently more spending per month compared to the online arm (**Table 4**). In the in-store arm, the median amount of voucher spent per month was the maximum value of $100 (IQR 98, 100) while a median of $91 (IQR 0, 100) per month was used in the online arm (p=0.0045).

**Table 4.**
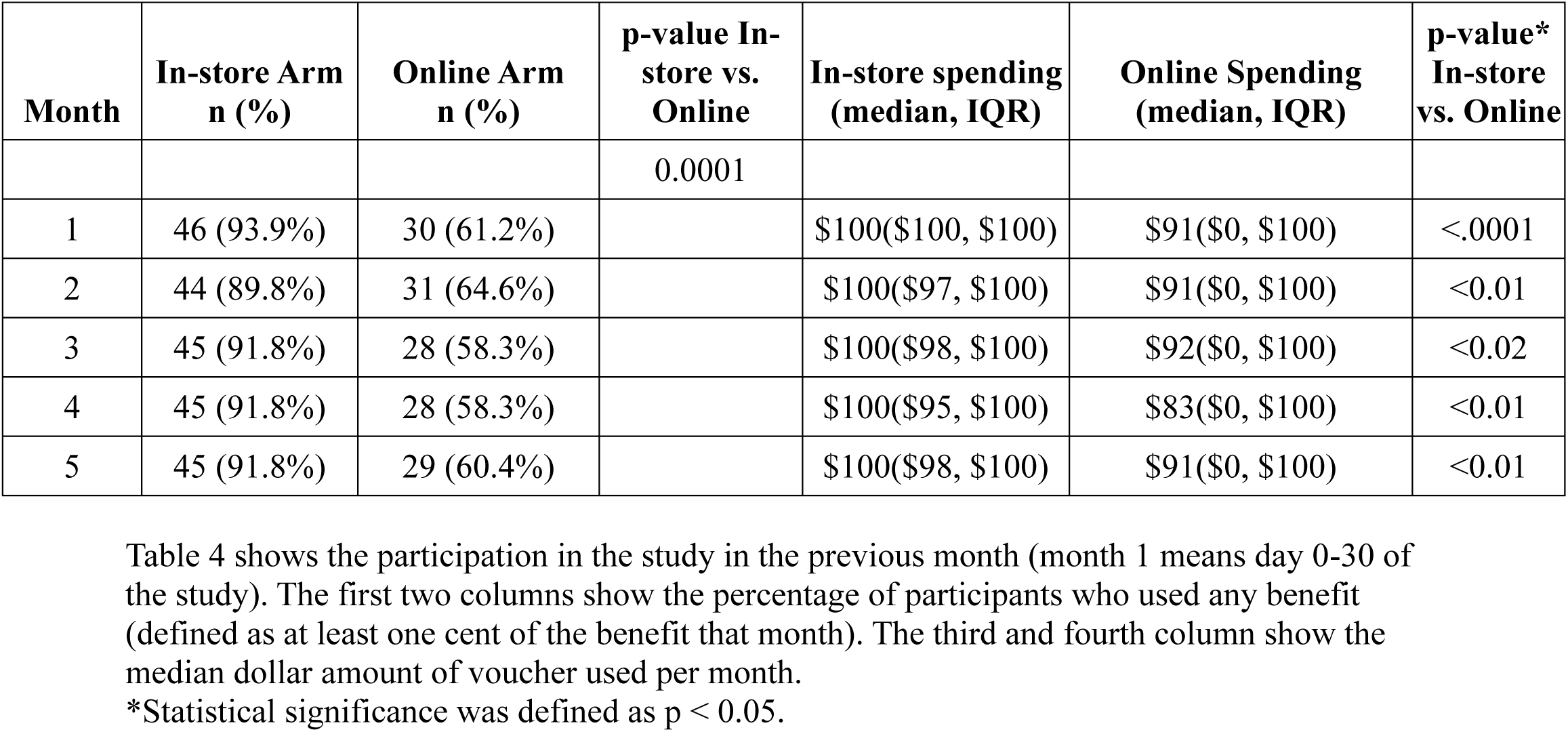
Percentage of Participants who used any benefit and median amount spent (in previous month)

| **Month** | **In-store Arm**<br>**n (%)** | **Online Arm**<br>**n (%)** | **p-value In-**<br>**store vs.**<br>**Online** | **In-store spending**<br>**(median, IQR)** | **Online Spending**<br>**(median, IQR)** | **p-value\***<br>**In-store**<br>**vs. Online** |
| --- | --- | --- | --- | --- | --- | --- |
|  |  |  | 0.0001 |  |  |  |
| 1 | 46 (93.9%) | 30 (61.2%) | | \$100(\$100, \$100) | \$91(\$0, \$100) | <.0001 |
| 2 | 44 (89.8%) | 31 (64.6%) | | \$100(\$97, \$100) | \$91(\$0, \$100) | <0.01 |
| 3 | 45 (91.8%) | 28 (58.3%) | | \$100(\$98, \$100) | \$92(\$0, \$100) | <0.02 |
| 4 | 45 (91.8%) | 28 (58.3%) | | \$100(\$95, \$100) | \$83(\$0, \$100) | <0.01 |
| 5 | 45 (91.8%) | 29 (60.4%) | | \$100(\$98, \$100) | \$91(\$0, \$100) | <0.01 |

Table 4 shows the participation in the study in the previous month (month 1 means day 0-30 of the study). The first two columns show the percentage of participants who used any benefit (defined as at least one cent of the benefit that month). The third and fourth column show the median dollar amount of voucher used per month.
\*Statistical significance was defined as $p < 0.05$ .

There was a bimodal distribution of money utilization in the online arm (**Figure 2**), meaning that a large portion of participants either spent $0 (31%) or all $100 (44%). Compared to online voucher users, online voucher non-users were more likely to be Caucasian (33% vs. 18%), employed (47% vs. 24%), be high income (20% vs 12%), and not have a college degree (80% vs. 70%).

**Figure 2.**
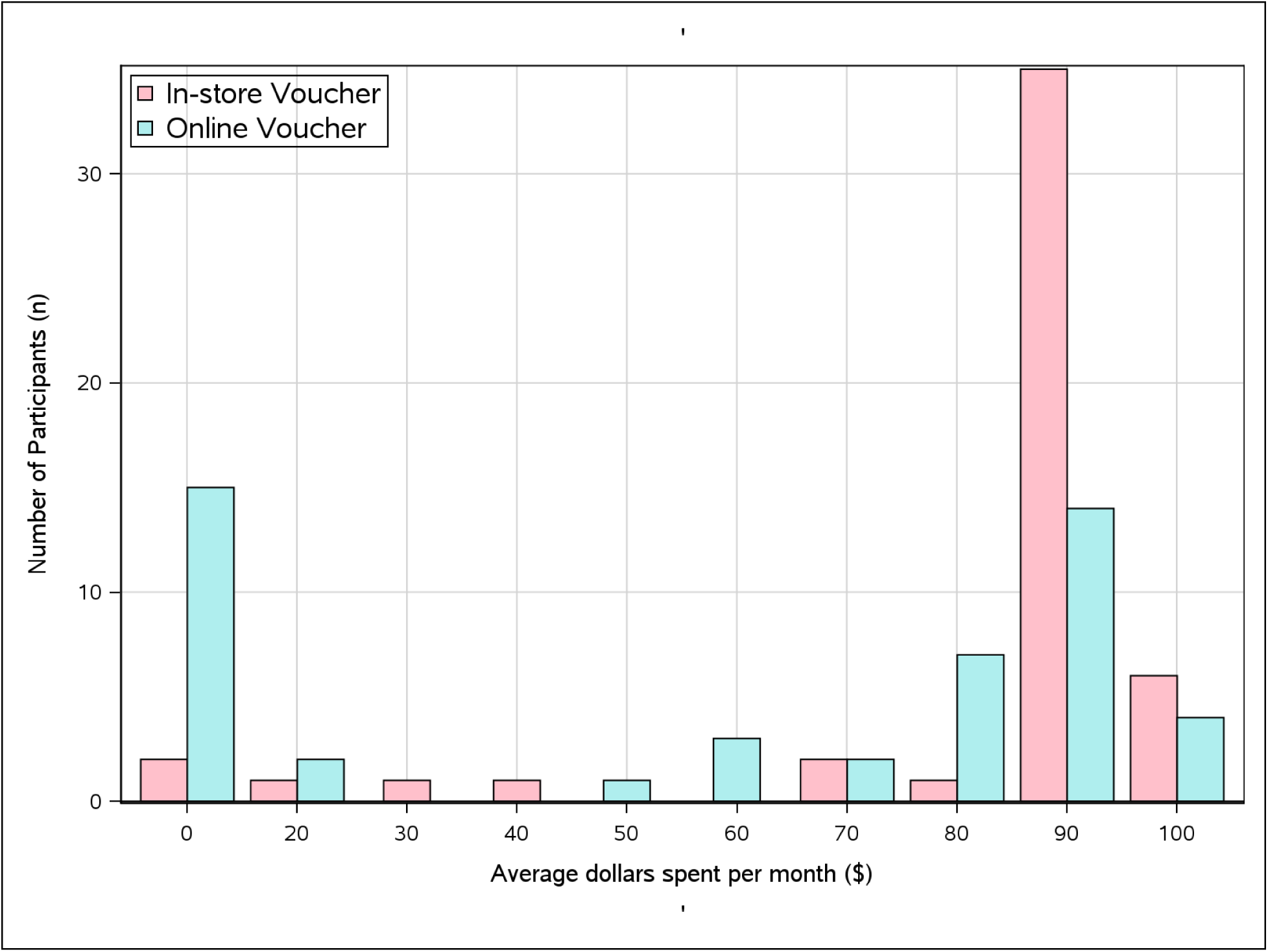
Histogram of Average Spending Per Month. Figure 3 is a histogram displaying the average number of dollars spent by each participant per month. The red bars are participants in the in-store voucher arm and the blue bars are participants in the online voucher arm.

The distribution of monetary spending was reliably collected for the online arm because of the established technical capabilities of the online platform which made this data readily and consistently available to the research team over the course of the study. In the online arm, the highest amount of voucher money over the course of the entire study was spent on meat (median $134 IQR [94, 165], **Table 6**) followed by fruits and vegetables (median $39, IQR [0, 74]). The lowest amount of voucher money was spent on vegetables alone (median $31, IQR [12, 45]). Because the same data infrastructure was not available through in-person grocer systems, voucher utilization is not reported for the in-person shopping group.

**Table 5.** Self-reported Use of Educational Material (in previous month)

| **Month** | **In-store Arm**<br>**n (%)** | **Online Arm**<br>**n (%)** | **p-value**<br>**In-store vs.**<br>**Online** |
| --- | --- | --- | --- |
|  |  |  | 0.0084 |
| 1 | 11 (22.4%) | 10 (20.4%) |  |
| 2 | 23 (46.9%) | 9 (18.8%) |  |
| 3 | 29 (59.2%) | 15 (31.3%) |  |
| 4 | 24 (49%) | 14 (29.2%) |  |
| 5 | 26 (53.1%) | 19 (39.6%) |  |

Table 5 shows the self-reported use of educational materials across randomization arms.

**Table 6.**
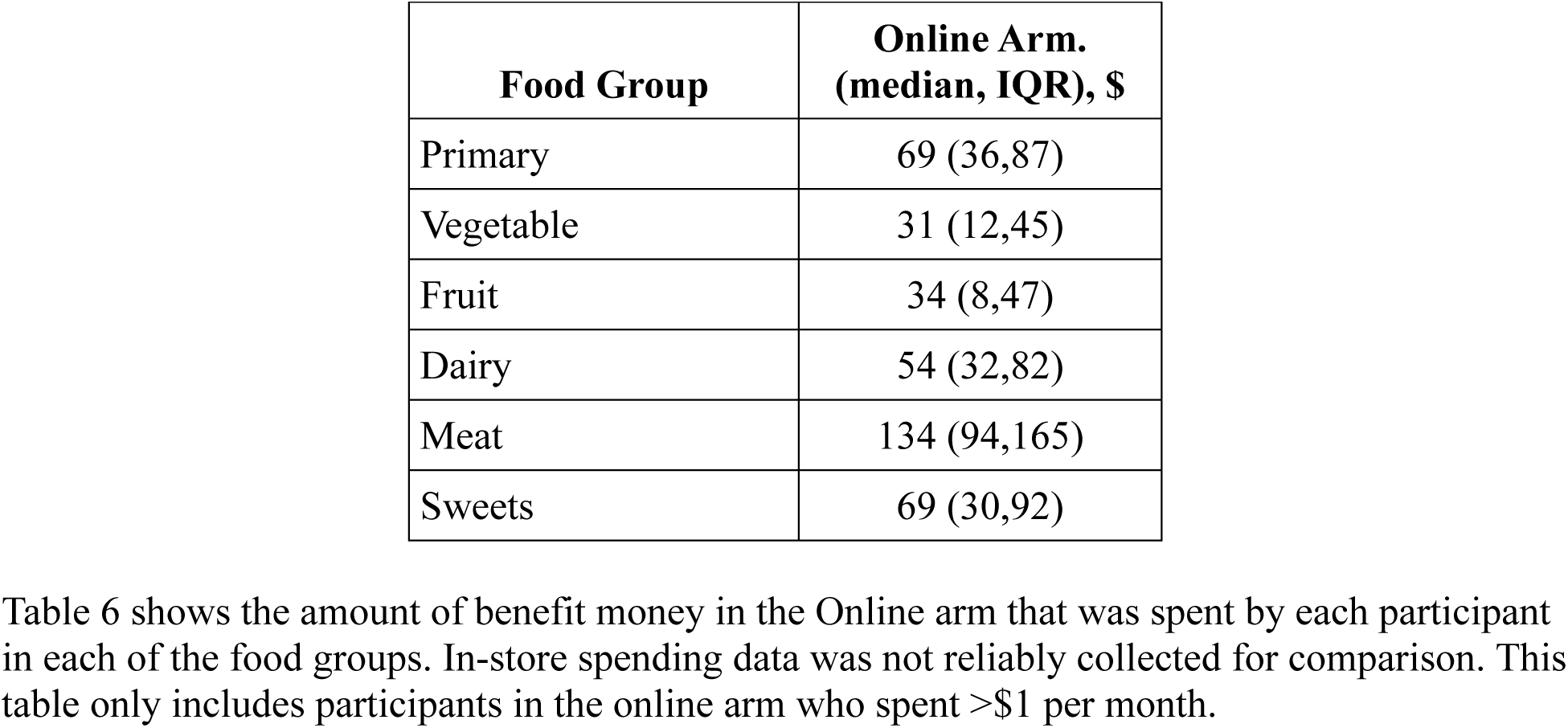
Voucher Spending in the Online Arm, over course of entire study.

| **Food Group** | **Online Arm.**<br>**(median, IQR), \$** |
| --- | --- |
| Primary | 69 (36,87) |
| Vegetable | 31 (12,45) |
| Fruit | 34 (8,47) |
| Dairy | 54 (32,82) |
| Meat | 134 (94,165) |
| Sweets | 69 (30,92) |

Table 6 shows the amount of benefit money in the Online arm that was spent by each participant in each of the food groups. In-store spending data was not reliably collected for comparison. This table only includes participants in the online arm who spent >\$1 per month.

### Self-reported Engagement with Educational Material

In the monthly FFQ, participants were asked if they read the educational materials in the previous month. Over 5 months, a significantly higher proportion of participants reported reading the educational materials in the in-store arm (22-59%) compared to the online arm (20-40%) (p=0.0084, **Table 5**).

## DISCUSSION

This randomized, pilot study compared the feasibility and effectiveness of two voucher-based incentives on food purchasing and consumption in patients with uncontrolled hypertension. The primary objective was to evaluate the feasibility of implementing these interventions and to identify trends in the effect of each intervention to inform a future, larger trial. We found that (1) there was no significant difference in self-reported food consumption patterns between an in-store and online voucher, (2) there was no significant change in average weight, BMI, systolic BP, or diastolic BP between or within arms, and (3) voucher utilization and engagement with educational materials was significantly higher in the in-store arm than the online arm.

Our findings build upon prior research evaluating the impact of financial vouchers on dietary behaviors. A meta-analysis performed by Hager *et al* examined the effects of 9 voucher-based interventions (paper vouchers or electronic cards ranging from $15-$300) across 22 sites (n=2064 adults and n=1817 children) on fruit and vegetable purchasing. After 6 months of intervention, they found that self-reported fruit and vegetable intake increased by 0.85 cups per day, Hgb A1c decreased by 0.29%, and systolic BP improved by 8 mmHg.^2^ Our study adds to this evidence by exploring if financially incentivized in-person versus online grocery shopping improved healthy food consumption. While no statistically significant differences were observed between the two groups, there was a trend toward increased vegetable consumption and reduced sweets consumption in the in-store voucher arm compared to the online arm. Similarly, within each voucher arm, there was also an overall trend towards increased vegetable consumption and decreased sweets consumption, suggesting that any type of voucher use may be associated with improved healthy food consumption.

Meta-analyses of prospective studies have demonstrated that a higher fruit and vegetable intake is associated with a lower risk of mortality and hypertension.^3,6^ Our study did not find a significant decrease in weight, BMI, systolic BP or diastolic BP. However, there was a trend towards decreased weight and BMI. Though this pilot study was not powered to detect statistical differences in food category consumption, the positive trend indicates that a larger study with longer follow-up period is warranted.

While we noted increased trends in vegetable consumption, our findings further highlight a substantial gap between dietary recommendations and current food and vegetable consumption levels. The U.S. Departments of Health and Human Services (DHHS) and Agriculture (USDA) recommend consuming 4-5 servings of fruits and vegetables per day, approximately 120-150 servings per month, as it is linked to a lower risk of mortality.^3,4^ In our study, the median monthly consumption of fruits and vegetables was 31.1 servings per month, and increased to 37.3 servings per month following intervention. These findings indicate that there are significant opportunities to enhance healthy eating behaviors among the urban population of New Orleans, Louisiana.

### Utilization of Vouchers and Educational Material

There was significantly higher voucher utilization in the in-store arm (>90% of participants) compared to the online arm (<64% of participants). This may reflect the greater familiarity and convenience of in-store shopping, which allowed participants to maintain their usual habits while using the voucher as a simple payment aid. In contrast, the online arm required downloading an app and adjusting to a new grocery ordering process, potentially creating barriers to use. Factors contributing to a technological divide such as internet access, age, education, income, and digital literacy should be considered when engaging populations for digitally driven food access programs.

A bimodal distribution was observed in online voucher use: 31% of participants did not use the voucher while over 44% used the full $100 value. This suggests that, despite initial usability challenges, those who adapted to the online platform maximally utilized the benefit. Thus, while online vouchers may present initial usability challenges, they could ultimately be an effective method for facilitating grocery purchases. Future online voucher programs may require targeted support to improve engagement among groups who exhibit digital challenges.

Participants in the in-store arm utilized the study’s educational materials more than the online arm. Although our study did not show a statistically significant improvement in healthy food purchasing, another pilot study found that combining vouchers with nutrition education increased healthy food consumption compared to vouchers alone.^8^ This highlights the potential impact of low-resource, self-directed educational material.

### Limitations and Lessons Learned

Barriers to the successful implementation of voucher-based FIM programs were identified through this pilot study. First, although voucher utilization was higher in the in-store arm, we were unable to obtain reliable itemized purchase data from the local grocery store, preventing a detailed analysis of expenditures across food categories (e.g., meat, dairy, vegetables). In contrast, the digital infrastructure of the online platform facilitated accurate and comprehensive data collection on participants’ purchases. Future partnerships with grocery stores should prioritize developing systems that enhance data accuracy, transfer, and accessibility for research.

Second, considerable effort was required to manually categorize grocery items into food groups (e.g., vegetables, fruit, dairy) to standardize for comparative analysis across grocery platforms. Streamlining this process with universal mapping dictionaries that can be shared across studies would facilitate efficient data collection and analysis in future FIM studies.

Third, the ability to restrict voucher-eligible purchases differed between platforms. The online platform allowed restrictions, preventing voucher use for alcohol and tobacco products, whereas no such limitations could be established for in-store vouchers. Consequently, a portion of in-store vouchers was spent on alcohol, which was not the intended use of the program. This highlights the need for mechanisms that ensure vouchers are restricted to healthy food purchases such as dietician curated or pre-selected options when implemented in store settings.

Fourth, some participants encountered challenges while using the digital platform to collect study forms and survey data. Additionally, the online grocery shopping platform required participants to provide an alternate form of payment to create an account which led to concerns among some participants who were uncomfortable sharing their financial information. These findings highlight the importance of assessing technological readiness and ensuring a user-friendly interface when implementing online components in similar programs.

Finally, while this study was underpowered to detect significant changes in food consumption, our findings suggest that local grocery store vouchers were more reliably utilized than mobile-based online vouchers. However, among participants who successfully adopted the online voucher, voucher utilization was maximized, with most spending the full amount. These findings suggest that future studies should prioritize targeted outreach efforts to populations that are less likely to use mobile-based grocery platforms such as individuals with technological barriers and low technological readiness to ensure equitable participation and effectiveness of interventions.

## CONCLUSION

This randomized, pilot study found no difference between in-store vouchers and online vouchers for consumption of fruits and vegetables. In both arms, there was a trend towards increased vegetable consumption and decreased sweets consumption over the 5 month intervention period, but these trends were not statistically significant. Future studies can improve implementation by streamlining data collection, refining voucher restrictions, standardizing food categorization, and providing targeted support for participant technological readiness.

## ABBREVIATIONS (in alphabetical order)

AHA: American Heart Association
BMI: Body Mass Index
BP: Blood Pressure
CI: Confidence Interval
EBT: Electronic Benefit Transfer
EHR: Electronic Health Records
FIM: Food is Medicine
FFQ: Food Frequency Questionnaire
GEE: Generalized Estimating Equations
GLM: General Linear Model
Hgb A1c: Hemoglobin A1c
HUGO: HUGO Health platform
IQR: Interquartile Range
LA: Louisiana
LCMC: Louisiana Children’s Medical Center
NHANES: National Health and Nutrition Examination Survey
PCORnet: Patient-Centered Outcomes Research Network
REDCap: Research Electronic Data Capture
SNAP: Supplemental Nutrition Assistance Program
USDA: U.S. Department of Agriculture
UMC: University Medical Center
UMCNO: University Medical Center New Orleans

## Data Availability

Data are available to the public here: https://zenodo.org/records/14776001

---

**Supplemental Table 1.**
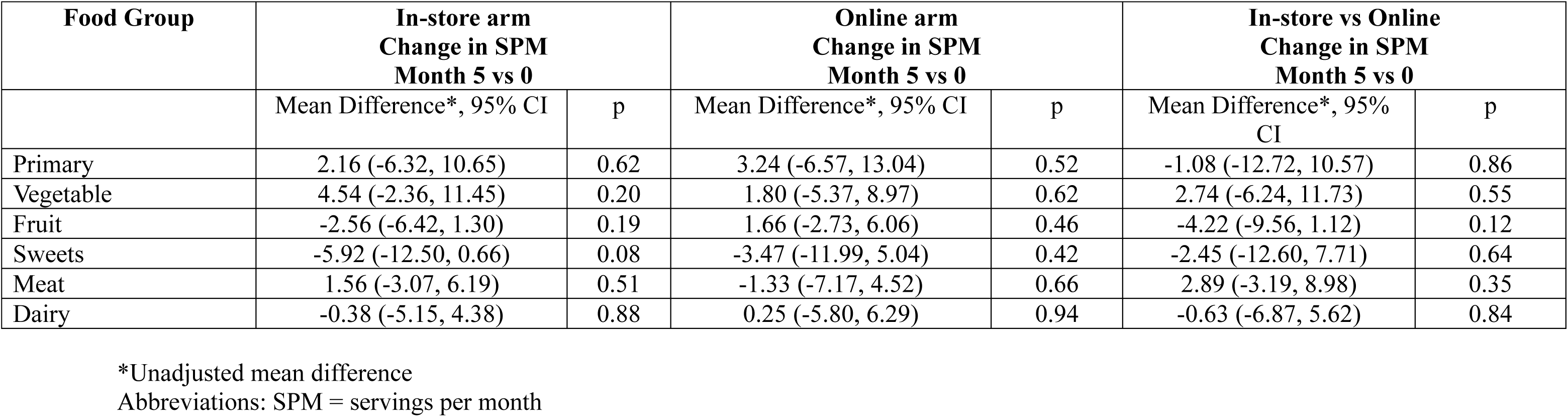
Unadjusted Mean Differences in Servings per Month Stratified by Randomization Arm and Food Group.

| Food Group | In-store arm<br>Change in SPM<br>Month 5 vs 0 |  | Online arm<br>Change in SPM<br>Month 5 vs 0 |  | In-store vs Online<br>Change in SPM<br>Month 5 vs 0 |  |
| --- | --- | --- | --- | --- | --- | --- |
|  | Mean Difference*, 95% CI | p | Mean Difference*, 95% CI | p | Mean Difference*, 95% CI | p |
| Primary | 2.16 (-6.32, 10.65) | 0.62 | 3.24 (-6.57, 13.04) | 0.52 | -1.08 (-12.72, 10.57) | 0.86 |
| Vegetable | 4.54 (-2.36, 11.45) | 0.20 | 1.80 (-5.37, 8.97) | 0.62 | 2.74 (-6.24, 11.73) | 0.55 |
| Fruit | -2.56 (-6.42, 1.30) | 0.19 | 1.66 (-2.73, 6.06) | 0.46 | -4.22 (-9.56, 1.12) | 0.12 |
| Sweets | -5.92 (-12.50, 0.66) | 0.08 | -3.47 (-11.99, 5.04) | 0.42 | -2.45 (-12.60, 7.71) | 0.64 |
| Meat | 1.56 (-3.07, 6.19) | 0.51 | -1.33 (-7.17, 4.52) | 0.66 | 2.89 (-3.19, 8.98) | 0.35 |
| Dairy | -0.38 (-5.15, 4.38) | 0.88 | 0.25 (-5.80, 6.29) | 0.94 | -0.63 (-6.87, 5.62) | 0.84 |

\*Unadjusted mean difference
Abbreviations: SPM = servings per month

**Supplemental Table 2.** Patient Characteristics by Online Usage.

|  | **Used Online**<br>**Voucher**<br>**(n=33)** | **Never Used Online**<br>**Voucher**<br>**(n=15)** |
| --- | --- | --- |
| Age (median, IQR) | 56 (53, 62) | 57 (51, 66) |
| Female (%) |  |  |
| Race (%) |  |  |
| Black | 70 | 53 |
| White | 18 | 33 |
| Multiple Races | 6 | 13 |
| Prefer not to answer | 6 | 0 |
| Ethnicity (%) |  |  |
| Hispanic | 3 | 7 |
| Non-Hispanic | 88 | 87 |
| Prefer not to answer | 9 | 7 |
| Income Category (%) |  |  |
| High Income,<br>>\$50,000 | 12 | 20 |
| Middle Income,<br>\$20,000-\$50,000 | 42 | 33 |
| Low Income,<br><\$20,000 | 42 | 40 |
| Unknown | 3 | 7 |
| Education Level (%) |  |  |
| College Graduate | 30 | 20 |
| Some College | 52 | 40 |
| High School | 15 | 40 |
| No Diploma | 3 | 0 |
| SNAP Benefit<br>Recipient (%) | 36 | 40 |
| Not employed (%) | 76 | 53 |
| Insurance Type (%) |  |  |
| Private | 9 | 20 |
| Public (Medicare,<br>Medicaid, VA) | 85 | 73 |
| Uninsured | 6 | 0 |
| Other |  |  |
| Number of people in<br>household (%) |  |  |
| 1 | 39 | 40 |
| 2 | 39 | 40 |
| 3 | 12 | 13 |
| ≥4 | 9 | 7 |
| BMI (kg/m<sup>2</sup>) | 35.4 (30.0, 38.1) | 33.4 (31.4, 37.6) |
| (median, IQR) |  |  |
| Average SBP in prior year (median, IQR) | 141.0 (137.0, 148.5) | 140.7 (136.0, 145.3) |
| Average DBP in prior year (median, IQR) | 80.8 (76.5, 86.5) | 85.7 (79.3, 89.6) |
| **Comorbidities (%)** |  |  |
| Cerebrovascular Disease | 9 | 0 |
| Diabetes with end-organ damage | 30 | 27 |
| Diabetes without end-organ damage | 33 | 40 |
| Heart failure | 15 | 0 |
| Malignant hypertension | 36 | 20 |
| Uncomplicated Hypertension | 79 | 80 |
| Chronic Lung Disease | 18 | 13 |
| Obesity | 39 | 53 |
| Peripheral vascular disease | 21 | 13 |

**Supplemental Table 3.** Overall Median Food Consumption Stratified by Food Group.

|  | Overall |  |  |
| --- | --- | --- | --- |
| Food Category | Median (IQR)<br>baseline | Median (IQR)<br>month 5 | % Change median<br>(IQR) |
| Primary (Vegetable +<br>Fruit) | 31.1 (14.2, 51.6) | 37.3 (28.6, 50.3) | 20.5 (-17.3, 93.0) |
| Vegetable | 19.3 (7.5, 32.9) | 26.1 (17.4, 32.9) | 27.7 (-9.7, 133.9) |
| Fruit | 8.7 (2.5, 15.2) | 15.2 (4.4, 15.2) | 0 (-42.9, 75.0) |
| Meat | 21.9 (13.7, 33.0) | 26.1 (15.5, 34.8) | 11.8 (-24.1, 75.0) |
| Dairy | 9.7 (6.0, 26.4) | 15.2 (8.7, 23.9) | 18.6 (-38.5, 113.8) |
| Sweets | 31.0 (17.3, 49.1) | 22.4 (8.5, 41.3) | -22.6 (-63.3, 21.4) |

This table shows the food consumption data stratified by food group. The food frequency is summarized using servings per month of each food group by patient self-report on the Food Frequency Questionnaire which was administered monthly.
Abbreviations: IQR = interquartile range

